# Evaluation of the diagnostic value of YiDiXie^™^-SS, YiDiXie^™^-HS and YiDiXie^™^-D in thyroid cancer

**DOI:** 10.1101/2024.07.03.24309931

**Authors:** Huimei Zhou, Yutong Wu, Peng Liao, Chen Sun, Zhenjian Ge, Wenkang Chen, Yingqi Li, Shengjie Lin, Pengwu Zhang, Wuping Wang, Siwei Chen, Xutai Li, Wei Li, Yongyan Cui, Jinsong He, Yongqing Lai

## Abstract

**Background:** Thyroid tumor, as an endocrine tumor with increasing incidence, causes a heavy economic burden. Thyroid ultrasound is widely used for screening or diagnosis of thyroid tumors. However, false-positive thyroid ultrasound results can lead to misdiagnosis and wrong puncture biopsy, while false-negative thyroid ultrasound results can lead to missed diagnosis and delayed treatment. There is an urgent need to find a convenient, cost-effective and non-invasive diagnostic method to reduce the false-positive and false-negative rates of thyroid ultrasound. The aim of the present study was to evaluate the diagnostic value of YiDiXie™-SS, YiDiXie™-HS and YiDiXie™-D in thyroid cancer.

**Patients and methods:** 843 subjects (malignant group, n=820; benign group, n=23) were finally included in this study. The remaining serum samples were collected and tested by YiDiXie ™ all-cancer detection kit. The sensitivity and specificity of YiDiXie™-SS were evaluated respectively.

**Results:** The sensitivity of YiDiXie™-SS was 98.5% (95% CI: 97.5% - 99.2%) and its specificity was 65.2% (95% CI: 44.9% - 81.2%). This means that YiDiXie ™ -SS has an extremely high sensitivity and relatively high specificity in thyroid tumors.YiDiXie™-HS has a sensitivity of 84.3% (95% CI: 81.6% - 86.6%) and a specificity of 87.0% (95% CI: 67.9% - 95.5%). This means that YiDiXie™-HS has high sensitivity and specificity in thyroid tumors.YiDiXie™-D has a sensitivity of 75.5% (95% CI: 72.4% - 78.3%) and a specificity of 95.7% (95% CI: 79.0% - 99.8%). This means that YiDiXie™-D has relatively high sensitivity and very high specificity in thyroid tumors.The sensitivity of YiDiXie™-SS in patients with positive thyroid ultrasound was 98.4% (95% CI: 97.3% - 99.1%) and the specificity was 64.3% (95% CI: 38.8% - 83.7%). This means that the application of YiDiXie ™ -SS reduced the false-positive thyroid ultrasound rate by 64.3% (95% CI: 38.8% - 83.7%) with essentially no increase in malignant tumor underdiagnosis.The sensitivity of YiDiXie™-HS in thyroid ultrasound-negative patients was 90.0% (95% CI: 79.9% - 95.3%) and the specificity 88.9% (95% CI: 56.9%). (95% CI: 56.5% - 99.4%). This means that the application of YiDiXie™-HS reduced the false negative rate of thyroid ultrasound by 90.0% (95% CI: 79.9% - 95.3%).YiDiXie ™-D has a sensitivity of 75.3% (95% CI: 72.1% - 78.2%) and a specificity of 92.9% (95% CI: 68.5% - 99.6%) in patients with positive thyroid ultrasound. This means that YiDiXie™-D reduces the rate of false-positive thyroid ultrasounds by 92.9% (95% CI: 68.5% - 99.6%). YiDiXie™-D has a sensitivity of 78.3% (95% CI: 66.4% - 86.9%) and a specificity of 100% (95% CI: 70.1% - 100%) in patients with negative thyroid ultrasounds. This means that YiDiXie™-D reduces the false-negative rate of thyroid ultrasound by 78.3% (95% CI: 66.4% - 86.9%) while maintaining high specificity.

**Conclusion:** YiDiXie™-SS has extremely high sensitivity and relatively high specificity in thyroid tumors.YiDiXie™-HS has high sensitivity and high specificity in thyroid tumors.YiDiXie ™ -D has relatively high sensitivity and extremely high specificity in thyroid tumors. YiDiXie™-SS significantly reduces thyroid ultrasound false-positive rates with essentially no increase in delayed treatment for thyroid cancer.YiDiXie ™ -HS significantly reduces thyroid ultrasound false-negative rates.YiDiXie™-D significantly reduces thyroid ultrasound false-positive rates or significantly reduces its false-negative rates while maintaining high specificity. YiDiXie™ tests have vital diagnostic value in thyroid cancer, and are expected to solve the problems of “high false-positive rate” and “high false-negative rate” of thyroid ultrasound.

**Clinical trial number:** ChiCTR2200066840.

## INTRODUCTION

Thyroid cancer is the most common endocrine malignancy and the fastest growing solid cancer in terms of incidence^1^. In 2020, there were 586,000 cases of thyroid cancer globally, which ranked 9th in terms of incidence among all malignant tumors^2^. In 2022, there were more than 821,000 cases of thyroid cancer globally in terms of total incidence, thyroid cancer ranks seventh among the most common cancers^3^. In recent years, there has been a rapid increase in incidence in many countries, dominated by papillary thyroid cancer^4^, which is largely attributed to the increasing use of ultrasound scanning and ultrasound-guided aspiration biopsies. Over 470,000 females and 90,000 males in the 12 countries counted may be overdiagnosed with thyroid cancer within 20 years^5^. Overdiagnosis due to ultrasound scanning leads to unnecessary fine-needle aspiration (FNA) biopsies, a dramatic increase in the diagnosis of thyroid cancer and subsequent surgery^6^. Overdiagnosis of thyroid cancer has prompted changes in clinical practice or guidelines in several countries^7-10^. As a result, thyroid tumors pose a serious threat to human health and a heavy economic burden.

Thyroid ultrasound produces not only a large number of overdiagnoses ^11-13^, but also a large number of false positive results. Studies have found that the risk of thyroid cancer in any patient who receives a subsequent diagnosis of nodules is only about 7-15%^14-18^. False-positive results for thyroid tumors imply misdiagnosis of benign disease as malignancy.Since puncture biopsy is usually taken in patients with positive results of ultrasound^10,19-20^. As a result, patients with false-positive thyroid ultrasound will have to bear unnecessary mental anguish, expensive tests, physical harm, and other adverse consequences. Based on the large and yearly increasing incidence of thyroid cancer^2-3^, false-positive results of thyroid ultrasound are becoming an increasing healthcare burden for mankind. Therefore, there is an urgent need to find a convenient, cost-effective and noninvasive method to reduce the false-positive rate of thyroid ultrasound.

Furthermore, thyroid ultrasound produces a large number of false negative results^14-18^. When thyroid ultrasound results are negative, patients are usually taken for observation and regular follow-up^10,19-20^. False-negative thyroid ultrasound results imply a missed diagnosis of thyroid cancer, which will likely lead to delayed treatment, progression of malignancy, and possibly even advanced stages. Patients will thus have to bear the adverse consequences of poor prognosis, expensive treatment, poor quality of life and short survival. Therefore, there is an urgent need to find a convenient, economical and noninvasive diagnostic method to reduce the false-negative rate of thyroid ultrasound.

Based on the detection of miRNAs in serum, Shenzhen KeRuiDa Health Technology Co., Ltd. has developed “YiDiXie ™ all-cancer test” (hereinafter referred to as the “YiDiXie™ test”)^21^. With only 200 milliliters of whole blood or 100 milliliters of serum, the test can detect multiple cancer types, enabling detection of cancer at home^21^. The “YiDiXie™ test” consists of three independent tests: YiDiXie ™-HS, YiDiXie™-SS and YiDiXie™-D^21^.

The purpose of this study is to evaluate the diagnostic value of YiDiXie™ tests in thyroid cancer.

## PATIENTS AND METHODS

### Study design

This work is part of the sub-study “Evaluating the diagnostic value of YiDiXie™ tests in multiple tumors” of the SZ-PILOT study (ChiCTR2200066840).

The SZ-PILOT study (ChiCTR2200066840) is a single-centre, prospective, observational study. Subjects who signed a pan-informed consent form for donation of remaining samples at the time of admission or physical examination were enrolled, and 0.5 ml of their remaining serum samples were collected for this study.

The study was blinded. Neither the laboratory personnel who performed YiDiXie ™ tests nor the KeRuiDa laboratory technicians who determined the results of YiDiXie ™ tests were aware of the subjects’ clinical information. The clinical experts who assessed the clinical information of the subjects were also unaware of the results of YiDiXie™ tests.

The study was approved by the Ethics Committee of Peking University Shenzhen Hospital and was conducted in accordance with the International Conference on Harmonization for “Good clinical practice guidelines” and the Declaration of Helsinki.

### Participants

The two groups were enrolled separately, with all subjects who met the inclusion criteria being consecutively included.

The study initially included inpatients with “suspected (solid or haematological) malignancy” who had signed a general informed consent form for donation of the remaining samples. Subjects with a postoperative pathological diagnosis of “malignant tumor” were included in the malignant group and those benign disease were included in the benign group. Subjects with ambiguous pathological findings were excluded from the study. Some of these malignant group subjects were used in our prior work^21^.

Subjects who failed the serum sample quality test prior to YiDiXie™ tests were excluded from the study. For further information on enrollment and exclusion, please see our prior work^21^.

### Sample collection, processing

The serum samples used in this study were obtained from serum left over after a normal clinic visit, without drawing additional blood. Approximately 0.5 ml of serum samples were collected from the remaining serum of subjects in the Medical Laboratory Department and stored at -80°C for subsequent use in YiDiXie™ tests.

### YiDiXie™ tests

YiDiXie™ tests is performed using the YiDiXie ™ all-cancer detection kit, an in vitro diagnostic kit developed and manufactured by Shenzhen KeRuiDa Health Technology Co., Ltd. It is an in vitro diagnostic kit for use in fluorescent quantitative PCR. It determines the presence of cancer in a subject’s body by measuring the expression levels of dozens of miRNA biomarkers in the serum. It predefines appropriate thresholds for each miRNA biomarker to ensure that each miRNA marker is highly specific, and integrates these independent assays in a concurrent testing format to significantly increase the sensitivity and maintain the specificity for broad-spectrum cancers.

YiDiXie ™ tests consists of three distinctly different tests: YiDiXie™-Highly Sensitive (YiDiXie™ -HS), YiDiXie ™ -Super Sensitive(YiDiXie ™ -SS) and YiDiXie™-Diagnosis (YiDiXie™-D). YiDiXie™-HS has been developed with sensitivity and specificity in mind. YiDiXie ™ -SS significantly increased the number of miRNA tests to achieve extremely high sensitivity for all clinical stages of all malignancy types. YiDiXie ™ -D dramatically increases the diagnostic threshold of individual miRNA tests to achieve very high specificity (very low false diagnosis rate) for all malignancy types.

Perform YiDiXie ™ tests according to the instructions of YiDiXie™ tests all-cancer detection kit. Refer to our prior work^21^ for detailed procedure.

The raw test results are analyzed by the laboratory technicians of Shenzhen KeRuiDa Health Technology Co., Ltd. and the results of YiDiXie ™ tests are determined to be “positive” or “negative”.

### Diagnosis of thyroid ultrasound

The diagnosis of thyroid ultrasound was determined according to the TI-RADS grade of the included cases if there was a TI-RADS grade. A TI-RADS grade of 4 or 5 was considered positive, and a TI-RADS grade of ≤ 3 was considered negative.

If there was no TI-RADS grade in the included cases, the results of the examination were considered “positive” or “negative” according to their diagnostic conclusions. Diagnoses that were positive, more positive, or tended to be malignant were considered “ positive “. If the diagnosis is positive, more positive or inclined to benign disease, or if the diagnosis is ambiguous, the test result is judged as “negative”.

### Clinical data collection

For this study, clinical, pathological, laboratory, and imaging data were extracted from the subjects’ hospitalized medical records or physical examination reports. All clinical staging was assessed by trained clinicians according to the AJCC staging manual (7th or 8th edition)^22-23^.

### Statistical analyses

For demographic and baseline characteristics, descriptive statistics were reported. For categorical variables, the number and percentage of subjects in each category were calculated; For continuous variables, the total number of subjects (n), mean, standard deviation (SD) or standard error (SE), median, first quartile (Q1), third quartile (Q3), minimum, and maximum values were calculated. 95% confidence intervals (CIs) for multiple indicators were calculated using the Wilson (score) method.

## RESULTS

### Participant disposition

843 study participants were involved in this research (n = 820 cases for the malignant group and 23 cases for the benign group). The 843 participants’ clinical and demographic details are listed in Table 1.

**Table 1.** Participants’ demographic and clinical manifestation.

|  | Malignant (n = 820) | Benign (n = 23) | Total (N = 843) |
| --- | --- | --- | --- |
| Age, years |  |  |  |
| Mean (SD) | 39.0 (10.48) | 46.6 (12.97) | 39.2 (10.62) |
| Median (Q1,Q3) | 37 (31, 45) | 52 (34, 58) | 37 (31, 46) |
| Min, max | 15, 73 | 22, 69 | 15, 73 |
| Age, group, n (%) |  |  |  |
| <50 | 663 (80.9) | 11 (47.8) | 674 (80.0) |
| ≥50 | 157 (19.1) | 12 (52.2) | 169 (20.0) |
| <65 | 808 (98.5) | 21 (91.3) | 829 (98.3) |
| ≥65 | 12 (1.5) | 2 (8.7) | 14 (1.7) |
| Sex, n (%) |  |  |  |
| Female | 617 (75.2) | 20 (87.0) | 637 (75.6) |
| Male | 203 (24.8) | 3 (13.0) | 206 (24.4) |
| Body mass index (kg/m<sup>2</sup>) |  |  |  |
| n | 809 | 23 | 832 |
| Mean (SD) | 23.3 (3.53) | 22.4 (2.71) | 23.3 (3.51) |
| Median (Q1,Q3) | 22.9 (20.7, 25.3) | 21.9 (20.4, 23.8) | 22.9 (20.7, 25.2) |
| Min, max | 14.6 39.0 | 16.1 27.4 | 14.6 39.0 |
| Body mass index category, n (%) |  |  |  |
| Underweight | 39 (4.1) | 1 (4.3) | 40 (4.7) |
| Benign | 455 (57.6) | 17 (73.9) | 472 (56.0) |
| Overweight | 240 (29.5) | 5 (21.7) | 245 (29.1) |
| Obese | 75 (7.9) | 0 (0) | 75 (8.9) |
| Missing | 11 (0.9) | 0 (0) | 11 (1.3) |
| AJCC clinical stage |  |  |  |
| Stage I | 796 (97.1) |  | 796 (97.1) |
| Stage II | 7 (0.9) |  | 7 (0.9) |
| Stage III | 17 (2.1) |  | 17 (2.1) |

Q1,Q3, first quartile, third quartile; SD, standard deviation.

In terms of clinical and demographic traits, the two study subject groups were similar (Table 1). The mean (standard deviation) age was 39.2 (10.62) years and 75.6% (637/843) were female.

### Diagnostic Performance of YiDiXie™-SS

As shown in Table 2, the sensitivity of YiDiXie™ -SS was 98.5% (95% CI: 97.5% - 99.2%), its specificity was 65.2% (95% CI: 44.9% - 81.2%).

**Table 2.**
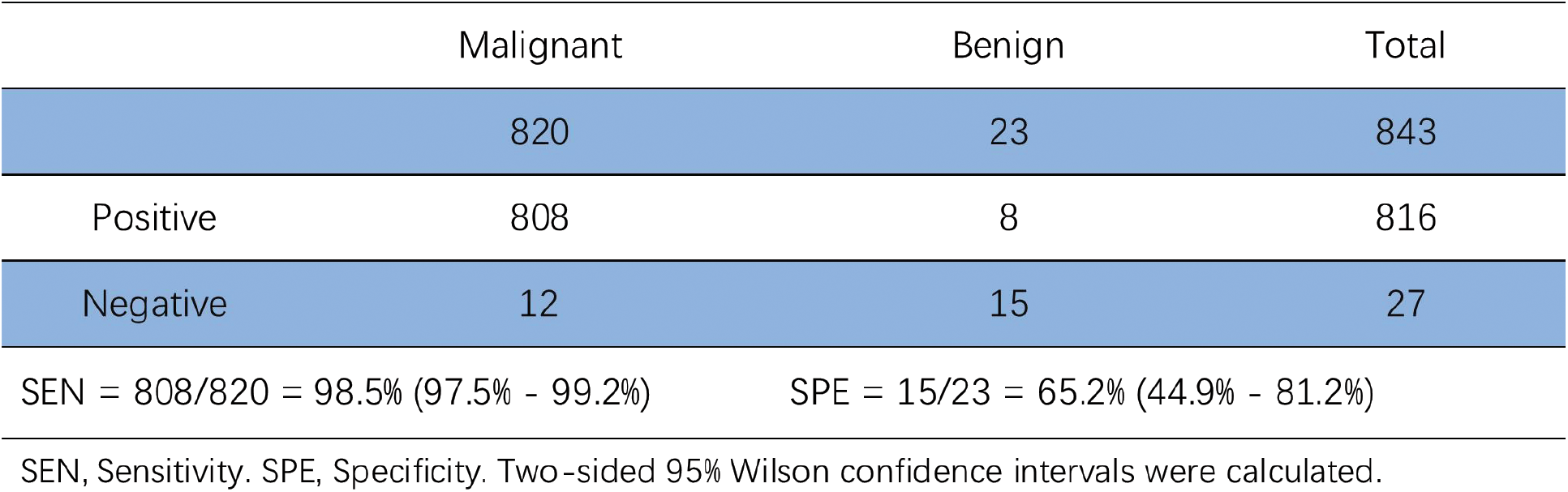
The performance of YiDiXie^™^ – SS.

|  | Malignant | Benign | Total |
| --- | --- | --- | --- |
|  | 820 | 23 | 843 |
| Positive | 808 | 8 | 816 |
| Negative | 12 | 15 | 27 |

SEN = $808/820 = 98.5\%$ (97.5% - 99.2%)
SPE = $15/23 = 65.2\%$ (44.9% - 81.2%)
SEN, Sensitivity. SPE, Specificity. Two-sided 95% Wilson confidence intervals were calculated.

### Diagnostic performance of YiDiXie™-HS

As shown in Table 3, the sensitivity of YiDiXie™ -HS was 84.3% (95% CI: 81.6% - 86.6%), its specificity was 87.0% (95% CI: 67.9% - 95.5%).

**Table 3.** The performance of YiDiXie^™^ - HS.

|  | Malignant | Benign | Total |
| --- | --- | --- | --- |
|  | 820 | 23 | 843 |
| Positive | 691 | 3 | 694 |
| Negative | 129 | 20 | 149 |

SEN = $691/820 = 84.3\%$ (81.6% - 86.6%)
SPE = $20/23 = 87.0\%$ (67.9% - 95.5%)
SEN, Sensitivity. SPE, Specificity. Two-sided 95% Wilson confidence intervals were calculated.

### Diagnostic performance of YiDiXie™-D

As shown in Table 4, the sensitivity of YiDiXie™ -D was 75.5% (95% CI: 72.4% - 78.3%) and its specificity was 95.7% (95% CI: 79.0% - 99.8%). This means that YiDiXie ™ -D has relatively high sensitivity and very high specificity in thyroid tumors..

**Table 4.** Performance of YiDiXie^™^ - D.

|  | Malignant | Benign | Total |
| --- | --- | --- | --- |
|  | 820 | 23 | 843 |
| Positive | 619 | 1 | 620 |
| Negative | 201 | 22 | 223 |

SEN = 619/820 = 75.5% (72.4% - 78.3%)
SPE = 22/23 = 95.7% (79.0% - 99.8%)
SEN, Sensitivity. SPE, Specificity. Two-sided 95% Wilson confidence intervals were calculated.

### Diagnostic performance of YiDiXie™-SS in thyroid ultrasound-positive patients

To address the challenge of high false-positive rate of thyroid ultrasound, YiDiXie™-SS was applied to thyroid ultrasound-positive patients.

As shown in Table 5, the sensitivity of YiDiXie™ -SS in thyroid ultrasound-positive patients was 98.4% (95% CI: 97.3% - 99.1%), and the specificity was 64.3% (95% CI: 38.8% - 83.7%). This means that the application of YiDiXie ™ -SS reduces the rate of thyroid ultrasound false positives by 64.3% (95% CI: 38.8% - 83.7%) with essentially no increase in malignant tumor underdiagnosis.

**Table 5.**
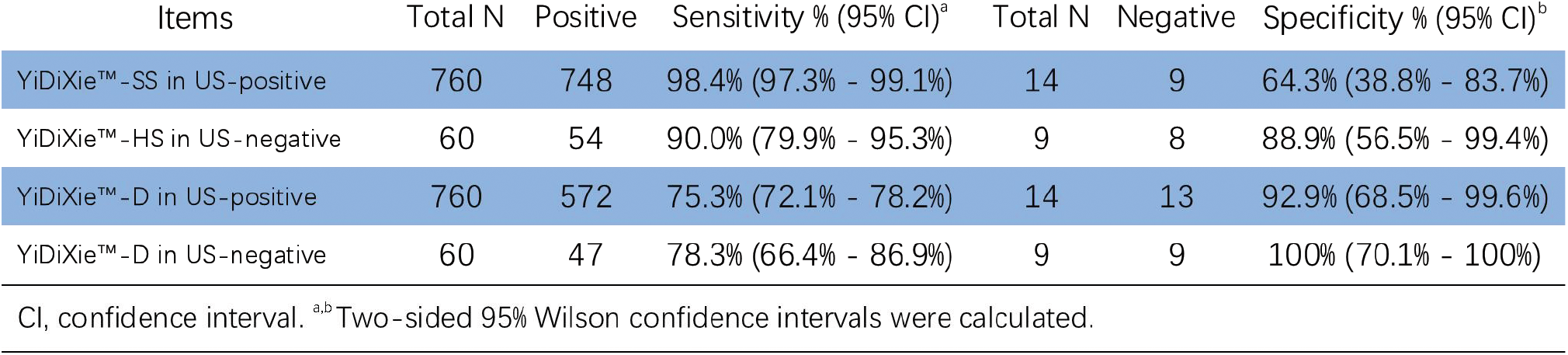
Performance of different Items.

### Diagnostic performance of YiDiXie™-HS in thyroid ultrasound-negative patients

To address the challenge of high false-negative rate of thyroid ultrasound, YiDiXie™ -HS was applied to thyroid ultrasound-negative patients.

As shown in Table 5, the sensitivity of YiDiXie™ -HS in thyroid ultrasound-negative patients was 90.0% (95% CI: 79.9% - 95.3%), and the specificity was 88.9% (95% CI: 56.5% - 99.4%). This means that the application of YiDiXie ™ -HS reduced the false negative rate of thyroid ultrasound by 90.0% (95% CI: 79.9% - 95.3%).

### Diagnostic performance of YiDiXie™-D in thyroid ultrasound-positive patients

To further reduce the rate of thyroid ultrasound false positivity, YiDiXie ™ -D was therefore applied to these patients.

As shown in Table 5, YiDiXie ™ -D had a sensitivity of 75.3% (95% CI: 72.1% - 78.2%) and its specificity was 92.9% (95% CI: 68.5% - 99.6%) in thyroid ultrasound positive patients. This means that YiDiXie™-D reduces the false positive rate of thyroid ultrasound by 92.9% (95% CI: 68.5% - 99.6%).

### Diagnostic performance of YiDiXie™-D in thyroid ultrasound-negative patients

In order to reduce the false-negative rate of thyroid ultrasound while maintaining high specificity, the more specific YiDiXie ™ -D was therefore applied to these patients.

As shown in Table 5, YiDiXie ™ -D has a sensitivity of 78.3% (95% CI: 66.4% - 86.9%) in thyroid ultrasound-negative patients and its specificity is 100% (95% CI: 70.1% - 100%). This means that YiDiXie ™ -D reduces the false-negative rate of thyroid ultrasound by 78.3% (95% CI: 66.4% - 86.9%) while maintaining high specificity.

## DISCUSSION

### Clinical significance of YiDiXie™-SS in thyroid ultrasound-positive patients

YiDiXie ™ tests consists of 3 tests with very different characteristics: YiDiXie™-HS, YiDiXie™-SS and YiDiXie ™ -D. Among them, YiDiXie ™ -HS combines high sensitivity and high specificity. YiDiXie ™ -SS has very high sensitivity for all malignant tumour types, but slightly lower specificity. YiDiXie™-D has very high specificity for all malignant tumour types, but lower sensitivity.

In patients with positive ultrasound scans for thyroid tumors, both sensitivity and specificity of further diagnostic methods are important. On the one hand, sensitivity is important. Lower sensitivity means a higher rate of false negatives. When the result of this diagnostic method is negative, the diagnosis usually ends for that patient. A higher false-negative rate means that more malignancies are missed, which will lead to delays in their treatment, progression of the malignancy, and possibly even development of advanced stages. Patients will thus have to bear the adverse consequences of poor prognosis, poor quality of life, and high cost of treatment.

On the other hand, specificity is important. Lower specificity means a higher rate of false positives. Thyroid tumors are usually puncture biopsied when the result of this diagnostic method is positive. Higher false positive rate means that more cases of benign cases are subjected to surgical resection or puncture biopsy. This undoubtedly substantially increases patients’ mental suffering, expensive surgery or examination costs, physical injuries, and other adverse consequences.

Therefore, the trade-off between sensitivity and specificity is essentially a trade-off between “fewer malignant tumors missed” and “fewer benign cases misdiagnosed”. In general, when a benign tumor is misdiagnosed as a malignant tumor, it is usually treated with simple tumor excision or puncture biopsy rather than radical resection. Thus, false positives for thyroid tumors do not lead to serious consequences in terms of organ loss. Thus, for thyroid tumors, it is far more important to have “fewer malignant tumors missed” than “fewer benign cases misdiagnosed”. Therefore, YiDiXie™-SS, with its very high sensitivity but slightly lower specificity, was chosen to reduce the false positive rate of ultrasound scans for thyroid tumors, rather than the highly sensitive and specific YiDiXie™-HS or YiDiXie™-D, which has very high specificity (very low false positive rate) but low sensitivity.

As shown in Table 5, YiDiXie ™ -SS had a sensitivity of 98.4% (95% CI: 97.3% - 99.1%) and a specificity of 64.3% (95% CI: 38.8% - 83.7%) in patients with positive thyroid ultrasound. These results suggest that YiDiXie ™ -SS reduces the false-positive rate of thyroid ultrasound by 64.3% (95% CI: 38.8% - 83.7%) with essentially no increase in malignant tumor underdiagnosis.

As mentioned earlier, a missed diagnosis of thyroid cancer means a delay in treatment, while a false-positive thyroid ultrasound means an unnecessary puncture biopsy. These results imply that YiDiXie™-SS drastically reduces the probability of erroneous puncture biopsy of benign thyroid cases with virtually no increase in malignant leakage diagnosis.

In other words, without basically increasing the delayed treatment of malignant tumors, YiDiXie ™ -SS substantially reduces the mental suffering, expensive examination costs, physical injuries, and other adverse consequences for patients with false-positive thyroid ultrasound. Hence, YiDiXie ™ -SS well fulfills the clinical demands and has important clinical significance and wide application prospects.

### Clinical significance of YiDiXie™-HS in thyroid ultrasound-negative patients

The sensitivity and specificity of further diagnostic methods are important in thyroid ultrasound-negative patients. Higher false-negative rates mean more thyroid cancers are missed. Higher false-positive rates mean more benign thyroid cases are misdiagnosed. In general, benign thyroid cases misdiagnosed as thyroid cancer usually undergo puncture biopsy, which is neither radical surgery nor affects the patient’s prognosis, and the cost of its puncture biopsy is much lower than the cost of advanced cancer treatment. Consequently, for patients with negative thyroid ultrasound, the “ risk of underdiagnosis of thyroid cancer “ is higher than the “ risk of misdiagnosis of benign thyroid cases”. Furthermore, the negative predictive value was higher in patients with negative thyroid ultrasound. Therefore, a higher false-positive and false-negative rate can lead to significant harm. Therefore, YiDiXie ™ -HS with high sensitivity and specificity was selected for reducing the false negative rate of thyroid ultrasound.

As shown in Table 5, YiDiXie ™ -HS had a sensitivity of 90.0% (95% CI: 79.9% - 95.3%) and a specificity of 88.9% (95% CI: 56.5% - 99.4%) in patients with negative thyroid ultrasound. This means that the application of YiDiXie™-HS reduced the false negative rate of thyroid ultrasound by 90.0% (95% CI: 79.9% - 95.3%).

The above results imply that YiDiXie ™ -HS substantially reduces the probability of missing malignant tumors with false-negative thyroid ultrasound results. Hence, YiDiXie™-HS well fulfills the clinical demands and has important clinical significance and wide application prospects.

### Clinical significance of YiDiXie™-D

In patients with thyroid tumors, YiDiXie ™ -D, with its relatively high sensitivity and very high specificity, can be used to further reduce the false-positive rate of thyroid ultrasound or to significantly reduce its false-negative rate while maintaining a high level of specificity.

As shown in Table 5, the sensitivity of YiDiXie™ -D in thyroid ultrasound-positive patients was 75.3% (95% CI: 72.1% - 78.2%), and its specificity was 92.9% (95% CI: 68.5% - 99.6%), while that of YiDiXie™ -D in thyroid ultrasound-negative patients was 78.3% (95% CI: 66.4% - 86.9%). 86.9%) in thyroid ultrasound-negative patients, and its specificity was 100% (95% CI: 70.1% - 100%). These results indicate that YiDiXie ™-D reduced thyroid ultrasound false positives by 92.9% (95% CI: 68.5% - 99.6%), or thyroid ultrasound false negatives by 78.3% (95% CI: 66.4% - 86.9%), respectively, while maintaining a high specificity.

The above results imply that YiDiXie ™ -D further reduces the risk of incorrect puncture biopsies of thyroid tumors. Therefore, YiDiXie™-D well meets the clinical needs and has important clinical significance and wide application prospects.

### YiDiXie™ tests are expected to solve the two problems of thyroid tumors

Firstly, YiDiXie ™ -SS significantly reduces the risk of misdiagnosis of benign thyroid cases as thyroid cancer. On the one hand, YiDiXie ™ -SS substantially reduces the probability of incorrect puncture biopsy in benign thyroid cases with essentially no increase in thyroid cancer missed diagnosis. As shown in Table 4, YiDiXie ™ -SS reduces the false-positive rate in 64.3% (95% CI: 38.8% - 83.7%; 9/14) of thyroid ultrasound-positive patients with essentially no increase in malignancy leakage. Accordingly, YiDiXie ™ -SS significantly reduces a range of adverse outcomes associated with unnecessary thyroid puncture biopsy with essentially no increase in delayed treatment of thyroid cancer.

On the other hand, YiDiXie ™ -SS reduces physician burdens and allows for rapid treatment of malignancies that might otherwise be postponed. When the ultrasound scan is positive, the patient usually undergoes puncture biopsy. The timely completion of these puncture biopsies is directly dependent on the number of physician. In many parts of the world, appointments of several months or even more than a year are available for thyroid puncture biopsy. It is inevitable that the treatment of malignant cases would be delayed. It is not uncommon for thyroid ultrasound-positive patients awaiting treatment to develop malignant progression or even distant metastases. As shown in Table 4, “YiDiXie™-SS reduces the false-positive rate of 64.3% (95% CI: 38.8% - 83.7%; 9/14) in patients with positive thyroid tumor ultrasound scans with essentially no increase in malignant tumor underdiagnosis. As a result, YiDiXie ™ -SS significantly relieves surgeons of unnecessary workload and facilitates the timely treatment of thyroid cancer or other diseases that would otherwise be delayed Secondly, YiDiXie ™ -HS significantly reduces the risk of underdiagnosis of thyroid cancer. When thyroid ultrasound is negative, the possibility of thyroid cancer is usually ruled out for the time being. The high rate of false-negative thyroid ultrasound results in a large number of thyroid cancer patients delaying treatment. As shown in Table 5, YiDiXie ™ -HS reduced the thyroid ultrasound false negative rate by 90.0% (95% CI: 79.9% - 95.3%; 54/60). Thus, YiDiXie ™ -HS substantially reduces the probability of missed malignancy diagnosis from false-negative thyroid ultrasound and facilitates timely diagnosis and treatment of thyroid cancer patients who would otherwise be delayed in treatment.

Thirdly, YiDiXie ™ -D is expected to further address the challenges of “high false positive rate” and “high false negative rate”. As shown in Table 5, YiDiXie ™ -D reduced thyroid ultrasound false positives by 92.9% (95% CI: 68.5% - 99.6%) or thyroid ultrasound false negatives by 78.3% (95% CI: 66.4% - 86.9%) while maintaining a high level of specificity. Thus, YiDiXie ™ -D further reduces the risk of incorrect puncture biopsies of thyroid tumors..

Finally, YiDiXie ™ tests enables “just-in-time” diagnosis of patients with positive ultrasound scans for thyroid tumors. On the one hand, YiDiXie ™ tests requires only a tiny amount of blood and allows patients to complete the diagnostic process non-invasively without having to leave their homes. Only 20 μ L of serum are required to complete a YiDiXie™ test, which is equivalent to approximately one drop of whole blood (one drop of whole blood is approximately 50 μ L, which yields 20-25 μ L of serum). Taking into account the pre-test sample quality assessment test and 2-3 repetitions, 0.2 ml of whole blood is sufficient for YiDiXie™ tests. The 0.2 ml of finger blood can be collected at home by the average patient using a finger blood collection needle without the need for venous blood collection by medical personnel, allowing the patient to complete the diagnostic process non-invasively without having to leave the patient’s home.

On the other hand, the diagnostic capacity of YiDiXie™ tests is nearly unlimited. Figure 1 shows the basic flow chart of YiDiXie™ test, from which it can be seen that the diagnostic capacity of YiDiXie ™ tests is virtually unlimited. YiDiXie ™ tests not only does not require a doctor or medical equipment, but also does not require medical personnel to collect blood.

**Figure 1.**
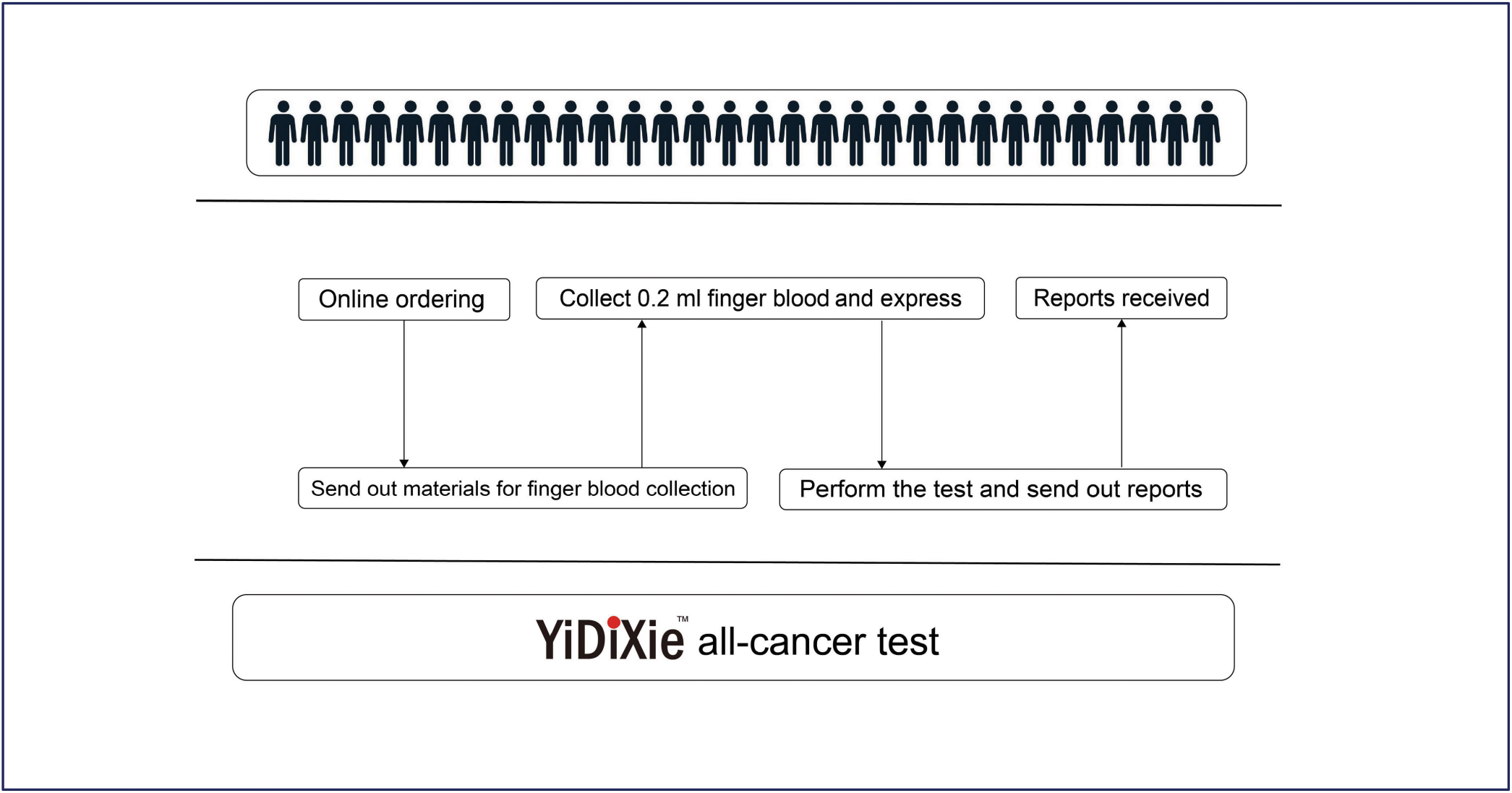
Basic flowchart of the “YiDiXie™ test”.

As a result, YiDiXie ™ tests is completely independent of the number of clinician and medical facilities, and has a virtually unlimited testing capacity. As a result, YiDiXie™ tests enables “just-in-time” diagnosis of patients with a positive ultrasound scan for thyroid tumors, without the need for patients to wait anxiously for an appointment.

In short, YiDiXie™ tests can play an important role in thyroid cancer, and are expected to solve the problems of “high false-positive rate of thyroid ultrasound” and “high false-negative rate of thyroid ultrasound”.

### Limitations of the study

Firstly, the number of cases in this study was small and future clinical studies with larger sample sizes are needed for further assessment.

Secondly, this study was a malignant tumor-benign tumor control study in inpatients, and future cohort studies of patients with thyroid tumor are needed for further assessment.

Finally, this study was a single-centre study, which may have led to some degree of bias in the results of this study. Future multi-centre studies are needed to further assess this.

## CONCLUSION

YiDiXie™-SS has extremely high sensitivity and relatively high specificity in thyroid tumors.YiDiXie™ -HS has high sensitivity and high specificity in thyroid tumors.YiDiXie ™ -D has relatively high sensitivity and extremely high specificity in thyroid tumors. YiDiXie ™ -SS significantly reduces thyroid ultrasound false-positive rates with essentially no increase in delayed treatment for thyroid cancer.YiDiXie ™ -HS significantly reduces thyroid ultrasound false-negative rates.YiDiXie ™ -D significantly reduces thyroid ultrasound false-positive rates or significantly reduces its false-negative rates while maintaining high specificity. YiDiXie ™ tests have vital diagnostic value in thyroid cancer, and are expected to solve the problems of “high false-positive rate” and “high false-negative rate” of thyroid ultrasound.

## Data Availability

All data produced in the present study are contained in the manuscript.

